# Provenance and funding of extremely cited biomedical papers published in 2003-2004, 2013-2014 and 2023-2024

**DOI:** 10.1101/2025.03.02.25323201

**Authors:** John P.A. Ioannidis

**Author notes:** Correspondence: John P.A. Ioannidis, MD, DSc, 3180 Porter Drive, Room A129, Stanford Research Park, Palo Alto CA 94304, USA.

## Abstract

**Importance:** It is important to monitor changes in biomedical literature and its funding. China has surpassed the USA in publications and, in some analyses, also in some impact indicators.

**Objective:** To evaluate changes over time in the profiles of the most highly cited biomedical papers.

**Design:** The 100 top-cited biomedical papers (based on Scopus) published in each of three time periods (2003-4, 2013-4, and 2023-4) were assessed for corresponding authors, types of publications represented, and funding sources, with emphasis on funding from the US National Institutes of Health (NIH) that has been traditionally considered the major funder of biomedical research.

**Setting:** Global.

**Participants:** not relevant

**Exposures:** not relevant

**Main outcome measures:** Provenance and funding sources.

**Main findings:** Corresponding authors from the USA decreased overtime (59/100 papers in 2003-4, 58/100 in 2013-4, 45/100 in 2023-4). China had corresponding authors in 0,1, and 4 top cited papers in the three time periods, respectively. There was a marked increase in consensus items (10/100 in 2003-4 versus 24/100 in 2023-4) and in reference statistics papers (1/100 in 2003-4, 10/100 in 2013-4, 11/100 in 2023-4). Reviews remained common among top cited papers, but almost always they were non-systematic. NIH funding was listed in 45/100, 50/100, and 23/100 papers in the three time periods, respectively. All other countries combined surpassed US public funding in 2023-4. Funding by NIH alone decreased sharply in the last decade (32/100, 28/100, and 2/100 in the three time periods, respectively). More commonly listed funding from non-profit organizations, societies, and institutions complemented the NIH funding decline. The first authors of 7/45 and the corresponding author(s) of 14/45 top cited USA-based papers of 2023-4 were listed as leaders of active NIH grants in RePORTER as of February 2025. Citation gaming became more obvious in 2023-4.

**Conclusions and relevance:** Overall, the USA remains a world leader regarding the most highly cited biomedical research and NIH funding retains a substantial presence among top cited papers. However, NIH influence has shrunk overall, and top cited papers funded exclusively by NIH have almost disappeared. Strengthening public funding is essential to secure research serves the common good.

---

The landscape of biomedical publications is changing over time in volume and provenance. The USA has long generated the majority of both overall publications and high-impact work. However, China now ranks first in numbers of annual publications, spends more on R&D and has more researchers (1–3). Some analyses have shown that in 2022 China outperformed the USA even in number of papers published in the 82 high-quality journals of the Nature Index (4); and excelled also in numbers of papers in top 1% of citations (5). Concurrently, major changes and challenges have emerged in research funding and the publication ecosystem. The US National Institutes of Health (NIH) have traditionally been the major public funder of biomedical research, but its budget has barely caught up with inflation and is currently threatened with major cuts (6). Concurrently, as publication numbers have increased rapidly, most investigators have struggled to secure funding from diverse sources. Publishers have large profit margins from a burgeoning publication market (7) that is subject to gaming forces on journal impact factors (8). Journals may also be willing to publish more papers that concentrate extremely high citations (9). Diverse gaming practices abound (10,11).

The current analysis aims to examine the most extremely cited biomedical papers published in 2003-2004, in 2013-2014 and most recently in 2023-2024. The following questions are asked: What is the provenance of these papers and what types of papers reached the extremes of citation distributions? Has the dominance of the USA indeed decreased and has China overtaken the USA in these citation extremes? What are the funding sources of these extremely cited papers? More specifically, has NIH continued to be the major funder of that work? Finally, might some of these extremely cited papers reflect citation gaming?

## Methods

### Search strategy and eligible papers

The analysis of Scopus data (12) is exploratory, and no protocol was pre-registered. No IRB approval was requested as it is a bibliometric analysis using public bibliometric data and there is also no reporting guideline for such studies. Only full papers were considered (Scopus categories of Articles, Reviews, and Conference Papers). Two periods were considered initially: full papers published in 2003-2004 and those published in 2023-2024. However, with stark differences observed, a third cohort of full papers published in 2013-2014 was added to probe whether changes were gradual. Searches for the first two cohorts were performed on January 24, 2025, and for the 2013-2014 cohort a month later. For each calendar period, all papers were ordered in decreasing number of citations received. Starting from the most-cited, publications were screened to identify those relevant to biomedicine and the top 100 most highly cited biomedical papers were thus retrieved for each of the time periods. Publications were deemed relevant to biomedicine if they belonged to any of the following Scopus subject areas: Medicine; Biochemistry, Genetics and Molecular Biology; Pharmacology, Toxicology and Pharmaceutics; Immunology and Microbiology; Neuroscience; Nursing; Health Professions; Veterinary; Dentistry. Papers catalogued in other subject areas by Scopus could be included only if they were deemed to have a major primary biomedical interest.

### Data extraction and definitions

For the three batches of 100 eligible papers each, meta-data were exported from Scopus on bibliographic information, citations, funding information, corresponding authors and their addresses. Moreover, full texts were downloaded to verify the Scopus extracted information and to complement missing information not captured by Scopus.

Country of origin was assigned based on the country of the corresponding author(s). The eligible papers were classified into the following types: Primary research: resources and methods (including new software, databases, methods [statistical, computational, or other] and tools); primary research: other; reviews; consensus items (including guidelines, recommendations, consensus documents, and definitions); and reference statistics for diseases and conditions. Funding was classified in the following categories: NIH; other US federal/public (e.g. NSF, DOE, CDC); other government/public from other countries; non-profit (e.g. Gates or Welcome Trust); for-profit (e.g. pharma or tech companies); societies (e.g. American Heart Association); and institutional (e.g. universities). When funding was listed from multiple categories, all of them were captured. Specifically for papers with listed NIH funding, they were also categorized as listing only NIH funding; or listing additional funders.

### Main outcomes

The main outcomes of interest are descriptive and include provenance (country of corresponding author), type of paper, type of funding, overall NIH funding, and NIH funding as sole funding source.

Furthermore, for the 2023-4 cohort, the first authors and the corresponding authors of papers with a USA corresponding author were assessed in NIH RePORTER (13) to see if they were listed as of February 2025 as the leader of any NIH grants.

Finally, it was assessed whether any of these highly cited papers had atypical features: >20% of the citations derived from a single journal; journal subject matter and/or title not representing the content of the article; or retraction (per Retraction Watch database) (14).

This project was aimed upfront to be descriptive without testing hypotheses with statistical tests.

## Results

### Scientific literature and identification of top cited biomedical papers

China was close to the USA in productivity by 2013-4 and had almost double the USA productivity in 2023-4 across the entire scientific literature, but the difference was more modest for the biomedical literature (Table 1). For all three time periods, biomedical papers accounted for most of the extremely cited papers across science. One had to screen 190, 185, and 162 top cited papers from 2003-4, 2013-4, and 2023-4, respectively, to identify 100 biomedical ones.

**Table 1.**
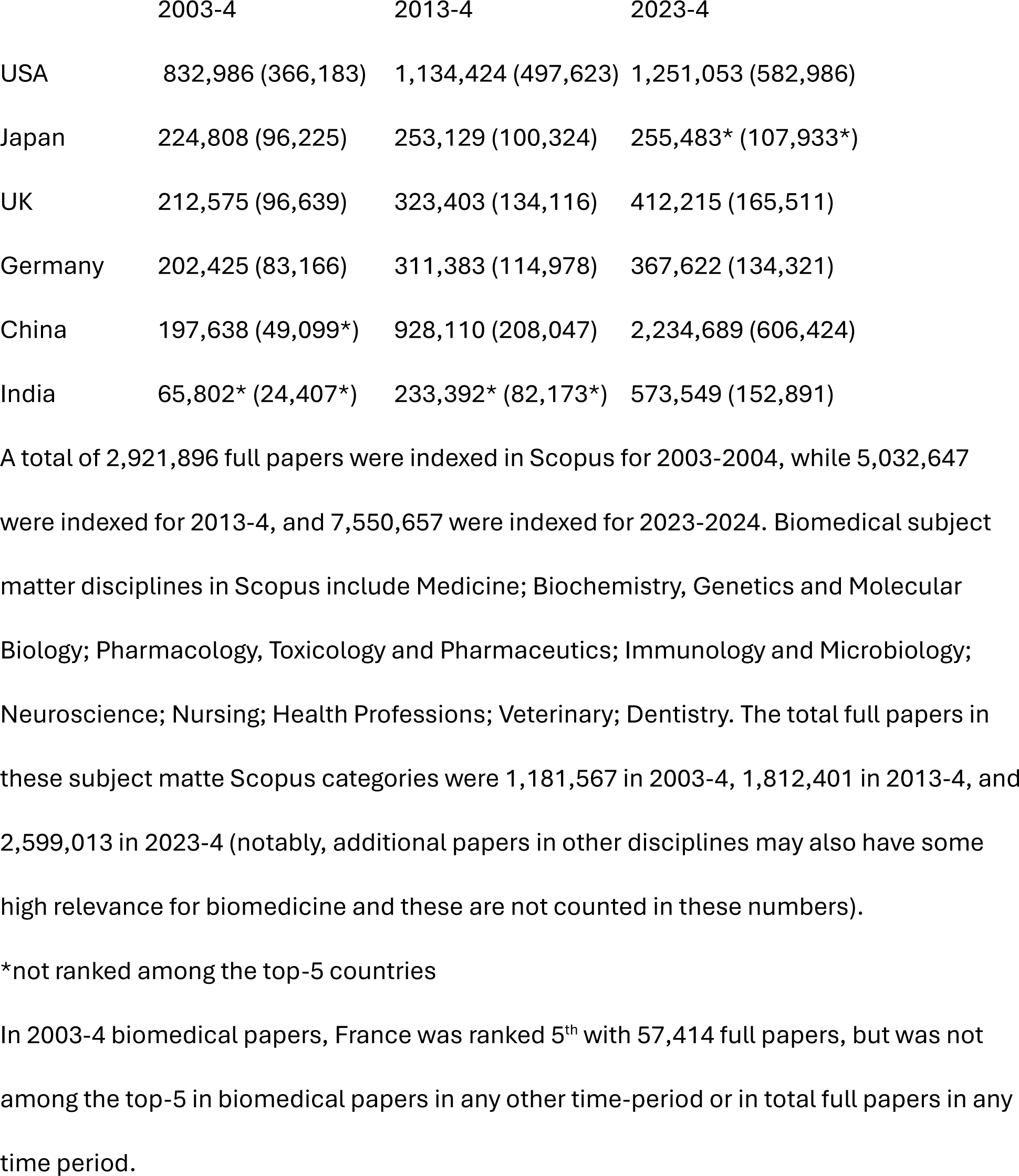
Top-5 countries with authors among all full papers (in parenthesis biomedical papers based on Scopus subject matter disciplines) published in 2003-4, 2013-4, and 2023-4 (per Scopus).

### Characteristics of the top cited biomedical papers (Table 2)

Details on the 100 most cited biomedical papers published in each of the three time periods appear in supplementary tables 1,2, and 3. A clear majority of papers in the 2003-4 cohort (59/100) and 2013-4 cohort (58/100) had USA corresponding authors, while in the 2023-4 cohort only 44/100 had USA corresponding authors. In the 2023-4 cohort there were 7 other countries with corresponding authors in at least 4 papers (6/7 European) versus only 3 such countries in earlier cohorts. 18 papers had corresponding authors from multiple countries in the 2023-4, while this was not seen in the 2003-4 cohort and had occurred only twice in the 2013-4 cohort. China had corresponding authors for 4 papers in 2023-4, 1 (in Hong-Kong) in 2013-4, and none in 2003-4. Besides China, 2 other non-high-income countries (India and Russia) also had 1 top cited paper each in 2013-2014, and 5 other non-high-income countries had corresponding authors in at least 1 paper in 2023-4 (India 2 papers, 1 paper each for Colombia, N. Cyprus, Jordan, Malaysia). Conversely, all papers in 2003-4 had corresponding authors from high-income countries.

**Table 2.** Characteristics of the 100 most cited biomedical papers published in 2003-4, 2013-4, and 2023-4.

|  | Publication year for the most cited papers |  |  |
| --- | --- | --- | --- |
|  | 2003-4 | 2013-4 | 2023-4 |
| Range of citations | 4983-48466 | 4403-53866 | 430-9785 |
| Corresponding author in USA* | 59 | 58 | 45 |
| Other countries with corresponding authors in $\geq 4$ papers | UK (8) | UK (10) | UK (16) |
|  | Germany (5) | Germany (8) | Germany (6) |
|  | Canada (4) | France (5) | Spain (6) |
|  |  |  | France (5) |
|  |  |  | Italy (4) |
|  |  |  | Switzerland (4) |
|  |  |  | China (4) |
| Corresponding authors in non-high income countries | 0 | 3 | 10 |
| Type of paper |  |  |  |
| Primary research** | 66 | 55 | 37 |
| Resources and methods | 34 | 30 | 17 |
| Other | 32 | 25 | 20 |
| Reviews | 23 | 20 | 28 |
| Non-systematic | 22 | 19 | 26 |
| Systematic | 1 | 1 | 2 |
| Consensus items*** | 10 | 15 | 24 |
| Reference statistics*** | 1 | 10 | 11 |

\*A few society-based papers did not list explicitly corresponding authors but were then allocated to the country of the society (unless the society was not based in a single country).
\*\*Some methodological papers are difficult to classify in the subgroup “Resources and methods” versus “Other”; only one of the two was chosen to avoid double-counting.
\*\*\*Consensus items and reference statistics may sometimes also use review methods (including systematic review methods), but they are not classified as Reviews, so as to keep the categories of types of paper mutually exclusive.

In 2003-4, two-thirds were primary research papers, equally split between “Resources and methods” work and other primary research. There were 23 reviews, of which only one aimed to be systematic;10 consensus items; and 1 reference statistics paper. The representation of primary research papers decreased gradually over time (55/100 in 2013-4 and 37/100 in 2023-4), relatively equally split between “Resources and methods” and other work. Reviews retained a considerable proportion, with almost all of them being non-systematic. Over a third of the top cited papers in the 2023-4 cohort were either consensus items (n=24) or reference statistics (n=11) with the increase in these types of papers noticed partly even in the 2013-2014 cohort.

### Funding of top cited papers

NIH was the leading single governmental funder in all three time cohorts, but the number of top cited papers with any listed NIH funding decreased in the last decade and became half in the 2023-4 cohort versus in the 2003-4 cohort or 2013-4 cohort (23 versus 45 or 50, respectively) (Table 3). When limited to primary research, NIH was listed as a funder in 15/37, 33/55, and 32/66 top cited papers, respectively, in these three periods.

**Table 3.** Funding of the 100 most cited biomedical papers published in 2003-4, 2013-4, and 2023-4

|  | Publication year for the most cited papers |  |  |
| --- | --- | --- | --- |
|  | 2003-4 | 2013-4 | 2023-4 |
| NIH | 45 | 50 | 23 |
| Other US federal/public | 4 | 9 | 4 |
| Other country public/government* | 29 | 43 | 37 |
| Non-profit | 13 | 21 | 24 |
| For-profit | 14 | 8 | 22 |
| Societies** | 6 | 9 | 25 |
| Institutional | 4 | 11 | 18 |
| None listed | 21 | 7 | 16 |

The sum for each cohort exceeds 100, because some papers listed more than one categories of funding.
\*includes international government-funded organizations such as International Agency for Research on Cancer (IARC), World Health Organization (WHO), and United Nations (UN).
\*\*when a paper was generated by one or more societies, these were considered to have funded the effort, even if funding was not explicitly listed.

In 2023-4, all other countries combined exceeded NIH and other US federal agencies in top cited papers. All European countries combined matched NIH in 2023-4, while China only funded 3 top cited papers. In 2023-4, there were large increases in numbers of top cited papers funded by non-profit organizations and those listing institutional support, and even more so for papers supported by societies. Some of this increase was seen already in 2013-4 and accentuated in the following decade.

In 2003-4, a substantive majority (32/45) of NIH-funded papers did not list any other funders and this remained true in 2013-4 (28/50) (Table 4). Conversely, there were only 2/23 NIH-funded papers that did not list any other funders in the 2023-4 cohort. Co-funding with non-profit organizations was common in all cohorts, but in 2023-4 there was also common co-funding with for-profit entities, public/government funding from other countries, and listed institutional support, which were less common in earlier time periods.

**Table 4.** Co-existence of other funders in the NIH-funded top cited papers

|  | Publication year for the most cited papers |  |  |
| --- | --- | --- | --- |
|  | 2003-4 | 2013-4 | 2023-4 |
|  | (n=45) | (n=50) | (n=23) |
| By presence of other funders |  |  |  |
| NIH alone | 32 | 28 | 2 |
| NIH and other listed funders | 13 | 22 | 21 |
| By type of other funders |  |  |  |
| Other US federal/public | 2 | 7 | 3 |
| Other country public/government* | 3 | 16 | 8 |
| Non-profit | 8 | 11 | 13 |
| For-profit | 4 | 5 | 7 |
| Societies** | 2 | 3 | 2 |
| Institutional | 2 | 4 | 7 |

\*includes international government-funded organizations such as IARC, WHO, and UN
\*\*when a paper was generated by one or more societies, these were considered to have funded the effort, even if funding was not explicitly listed.

### Current NIH funding for first and corresponding authors

Of the 45 top cited papers of 2023-4 with USA-based corresponding authors, in 7 (16%) the first author was listed in RePORTER as leading some currently active NIH grant(s). The same was true for at least one of the corresponding authors in 14 papers (31%).

### Atypical papers

One paper in 2023 was a guideline for reporting surgical case reports and >98% of its citations had been received by the publishing journal which indeed publishes many surgical case reports. Another paper in 2023 was published in a Chemistry journal, and the title suggested that it was providing cancer statistics and projections to 2030, but the content was a review of diverse cancers without substantive epidemiological projections.

Apparently, the paper was heavily cited by authors who mistook it for other regularly updated papers on cancer statistics. No top cited papers in any time cohort had been retracted.

## Discussion

Biomedical research represents the majority of the most highly cited scientific literature and USA-based authors continue to dominate the most influential scientific literature in biomedicine, despite modest decline in the last decade. In contrast to previous work focused on publications in prestigious journals (4) or the top 1% of citations (5), where China apparently outpaced the USA, the current analysis shows that China still has negligible presence among the 100 most highly cited recent biomedical papers. NIH continues to be commonly listed as funder. Nevertheless, in recent papers, NIH is listed only about half that often compared with one or two decades ago. More prominently, in the recent cohort of extremely highly cited papers, NIH is almost never listed as the only funder, in stark contrast with just a decade ago.

Non-profit organizations, societies, and institutional support are increasingly listed as funders in top cited papers. Recent studies tend to have a wider range of acknowledged funders. Perhaps reporting of funders has become more detailed, on average, over time.

Investigators may take more care to acknowledge support and/or journals may be more explicit in funding disclosures (14). Some institutional support is likely to have been and continue to be common in all work from universities and research institutions. However, in the past this has typically not been acknowledged explicitly. Public funding may also have misclassification. Some investigators may not have listed their NIH funding, even if relevant to the work. Conversely, some other investigators may have listed NIH funding even if irrelevant to the published work. The latter pattern may become more frequent if investigators are increasingly pressured to demonstrate productivity to sponsors.

While other funders may cover some of the gap left by public research funding (15), public funding of the form provided by NIH is important to strengthen. Funding cuts may exacerbate pressure to game the system. The large majority of the first authors or even the corresponding authors (who may be more senior, on average) of the most recent USA-based extremely highly cited biomedical papers do not lead any currently active NIH grants. This is consistent with earlier analyses of NIH funding (16). More rather than less funding is therefore needed. Still, evidence-based efforts at research reform are welcome, including assessment of review, evaluation and funding distribution methods (17–19).

The top cited papers of 2023-4 show an increasing footprint of the pressure to boost impact metrics. The advent of massive publications has been associated with perverse incentives and even an increasing rate of fraud (20,21). This is particularly true for China (22,23). Outright fraud may be distinctly uncommon in the sample of extremely cited papers analyzed here, but gaming is likely. E.g., the share of consensus items has increased markedly compared with earlier periods. Guidelines and other consensus documents have seen major increases in production (24). Consensus documents can be very important, but the methodological rigor, and protection from conflicts for most of them is questionable (25,26). Some journals have boosted their impact factors by engaging fervently in production of consensus documents. Another type of publication that achieves similar goals is the release of reference statistics. They can also be valuable, but they also provide routine citations for introduction sections. There is less methodological research on the validity of these exercises, but some early concerns have been raised even for the most rigorous among them (27).

The popularity of such papers may also lead to the publication of manuscripts that mimic their titles and thus share citation success. Authors usually do not read the papers they cite (28) and titles may mislead about the content. Another atypical situation documented here is the publication of a reporting guidance paper that is widely cited in articles appearing in the publishing journal. Finally, the considerable prevalence of reviews among the extremely cited publications is unsurprising. Reviews can attract many citations and often reflect the culmination or integration of important work and insightful syntheses.

A worrisome pattern is the pervasive dearth of systematic approaches among extremely cited reviews. Several biases might have been curtailed with systematic methods (29,30). In some fields, especially in basic science, non-systematic reviews still draw a lot of attention, and they may be thought to provide a broader overview of the field. However, higher adoption of systematic review methods would improve also basic science (31).

Beyond highly cited articles, other indicators may be examined to obtain a comprehensive view of the evolution and current state of the research ecosystem in the USA, China, and globally. China has outpaced the USA not only in number of overall publications, but also in patent applications (1,619,268 versus 594,340 in 2023), patent grants (798,347 versus 323,410 in 2023) (32), and new business applications (24.8 million in the first 9 months of 2023 (33) versus 5.5 million in the entire 2023 (34), USA publications have a higher rate of international collaboration than China (40% versus 22% in 2023), but many other countries have even higher rates of international collaboration than the USA (35). The decrease over time in the role of NIH is congruent with the decrease in the proportion of R&D funding from government sources in the USA over time. In the USA, government sources accounted for 20% of R&D in 2021 (latest available data, ref. 36 https://ncses.nsf.gov/pubs/nsb20246/cross-national-comparisons-of-r-d-performance), while they represented 32% in 2010 (37) and 57% in the 1970s (38). In 2025, there have been major threats for further sharp reductions in federal funding and increased uncertainty. Conversely, the proportion of R&D with government source is higher in China and the overall R&D expenditure in China was very close to that of the USA in 2021 and may have fully matched the USA by now, if the long-term trends continued in the last 4 years (39).

Some limitations should be acknowledged. First, citations are not equated with disruptive innovation. However, it is difficult to measure disruption objectively, even more so in recent work. There is some worrisome (40) but debated (41) evidence for substantial recent decline in disruptiveness. Second, the citation potential of recently published papers has not been fully expressed yet. Moreover, given the substantially longer time available to be cited, papers published early in 2023 enjoy major advantages to reach the top cited list than those published in late 2024. However, relative timing of publication within 2023-4 is unlikely to be affected by country of provenance, type of paper or funding pattern. There is no evidence that papers from a specific country, type, or funding are substantially more likely to start acquiring massive citations late, while relatively unnoticed early on (42). Such “sleeping beauties” are rather uncommon (42).

Acknowledging these caveats, the analyzed papers offer key referential landmarks for large, prolific and influential fields. This applies not only to traditional primary basic science research, but also to centralized resources, new methods, high-impact randomized trials, and even consensus statements, statistics, and reviews. In the current landscape, while the US remains a key player, its influence has diminished. The decrease is even more prominent for the presence of NIH funding and very prominent for solitary NIH funding. Strong public support of science is indispensable to ensure that research retains an orientation towards enhancing the public good. With threats for major cuts in government funding in 2025, it is important to avoid science becoming a political battlefield and secure bipartisan support for biomedical research.

## Data sharing statement

All data are in the manuscript and in the supplementary tables.

## Contributions

JPAI had the idea, collected the data, analyzed the data, and wrote the paper.

## Data Availability

All data are in the manuscript and in the supplementary tables

## SUPPLEMENTARY TABLES

**Supplementary Table 1.** 100 top cited biomedical papers published in 2003-4

| Title | Type of paper | Year | Source | Cited | Country | Classified funding |
| --- | --- | --- | --- | --- | --- | --- |
| Measuring inconsistency in meta-analyses | Resource | 2003 | BMJ | 48466 | UK | UK |
| MUSCLE: Multiple sequence alignment with high accuracy and high throughput | Resource | 2004 | Nucleic Acids Research | 35453 | USA | European |
| UCSF Chimera - A visualization system for exploratory research and analysis | Resource | 2004 | Journal of Computational Chemistry | 34953 | USA | NIH |
| Cytoscape: A software Environment for integrated models of biomolecular interaction networks | Resource | 2003 | Genome Research | 34551 | USA | NIH |
| MicroRNAs: Genomics, Biogenesis, Mechanism, and Function | Resource | 2004 | Cell | 31906 | USA | NIH/non-profit |
| MrBayes 3: Bayesian phylogenetic inference under mixed models | Resource | 2003 | Bioinformatics | 27150 | Sweden | NSF/Sweden |
| Coot: Model-building tools for molecular graphics | Resource | 2004 | Acta Crystallographica Section D | 25964 | UK | UK/societies |
| Classification of surgical complications: A new proposal with evaluation in a cohort of 6336 patients and results of a survey | Resource | 2004 | Annals of Surgery | 25433 | Switzerland | None |
| EEGLAB: An open source toolbox for analysis of single-trial EEG dynamics including independent component analysis | Resource | 2004 | Journal of Neuroscience Methods | 17538 | USA | NIH/non-profit/institution |
| The seventh report of the joint national committee on prevention, detection, evaluation, and treatment of high blood pressure: The JNC 7 report | Consensus | 2003 | JAMA | 17348 | USA | NIH |
| International physical activity questionnaire: 12-Country reliability and validity | Resource | 2003 | Medicine and Science in Sports and Exercise | 15013 | Australia | None |
| Development and testing of a general Amber force field | Resource | 2004 | Journal of Computational Chemistry | 14870 | USA | NIH |
| Qualitative content analysis in nursing research: Concepts, procedures and measures to achieve trustworthiness | Resource | 2004 | Nurse Education Today | 13264 | Sweden | None |
| Global Prevalence of Diabetes: Estimates for the year 2000 and projections for 2030 | Reference statistics | 2004 | Diabetes Care | 12676 | UK | None |
| Seventh report of the Joint National Committee on Prevention, Detection, Evaluation, and Treatment of High Blood Pressure | Consensus | 2003 | Hypertension | 11236 | USA | NIH |
| Mfold web server for nucleic acid folding and hybridization prediction | Resource | 2003 | Nucleic Acids Research | 11185 | USA | NIH |
| MEGA3: Integrated software for Molecular Evolutionary Genetics Analysis and sequence alignment. | Resource | 2004 | Briefings in bioinformatics | 10724 | USA | NIH/NSF/non-profit |
| Advances in functional and structural MR image analysis and implementation as FSL | Resource | 2004 | NeuroImage | 10689 | UK | UK/for-profit/non-profit |
| Activating Mutations in the Epidermal Growth Factor Receptor Underlying Responsiveness of Non-Small-Cell Lung Cancer to Gefitinib | Article | 2004 | NEJM | 10509 | USA | NIH |
| Prejudice, Social Stress, and Mental Health in Lesbian, Gay, and Bisexual Populations: Conceptual Issues and Research Evidence | Article | 2003 | Psychological Bulletin | 10327 | USA | NIH |
| Bioconductor: open software development for computational biology and bioinformatics. | Resource | 2004 | Genome biology | 9996 | USA | None |
| Chronic kidney disease and the risks of death, cardiovascular events, and hospitalization | Article | 2004 | NEJM | 9949 | USA | NIH |
| The functions of animal microRNAs | Review | 2004 | Nature | 9717 | USA | NIH |
| Bevacizumab plus irinotecan, fluorouracil, and leucovorin for metastatic colorectal cancer | Article | 2004 | NEJM | 9669 | USA | NIH |
| Linear models and empirical bayes methods for assessing differential expression in microarray experiments | Resource | 2004 | Statistical Applications in Genetics and | 9650 | Australia | Australia |
|  |  |  | Molecular<br>Biology |  |  |  |
| WebLogo: A sequence logo generator | Resource | 2004 | Genome<br>Research | 9649 | USA | NIH |
| Appropriate body-mass index for Asian<br>populations and its implications for policy and<br>intervention strategies | Article | 2004 | Lancet | 9541 | Switzerland | Japan/Singapore |
| Effect of potentially modifiable risk factors<br>associated with myocardial infarction in 52<br>countries (the INTERHEART study): Case-control<br>study | Article | 2004 | Lancet | 9454 | Canada | Canada/for-profit/non-profit |
| APE: Analyses of phylogenetics and evolution in R<br>language | Resource | 2004 | Bioinformatics | 9296 | France | Germany |
| Prospective identification of tumorigenic breast<br>cancer cells | Article | 2003 | PNAS | 8961 | USA | NIH |
| EGFR mutations in lung, cancer: Correlation with<br>clinical response to gefitinib therapy | Article | 2004 | Science | 8893 | USA | NIH |
| Exploration, normalization, and summaries of high<br>density oligonucleotide array probe level data. | Resource | 2003 | Biostatistics | 8770 | USA | NIH |
| The Benefits of Being Present: Mindfulness and Its<br>Role in Psychological Well-Being | Review | 2003 | Journal of<br>Personality and<br>Social<br>Psychology | 8530 | USA | NIH/societies/non-profit/Canada |
| Essential oils: Their antibacterial properties and<br>potential applications in foods - A review | Review | 2004 | International<br>Journal of Food<br>Microbiology | 8444 | Netherlands | None |
| The biology of VEGF and its receptors | Review | 2003 | Nature Medicine | 8424 | USA | None |
| Staging of brain pathology related to sporadic<br>Parkinson's disease | Article | 2003 | Neurobiology of<br>Aging | 8338 | Germany | Germany/Australia |
| Obesity is associated with macrophage<br>accumulation in adipose tissue | Article | 2003 | Journal of<br>Clinical<br>Investigation | 8216 | USA | NIH |
| The International Classification of Headache<br>Disorders: 2nd edition. | Consensus | 2004 | Cephalalgia | 8168 | Denmark | None |
| DAVID: Database for Annotation, Visualization, and Integrated Discovery. | Resource | 2003 | Genome biology | 8052 | USA | NIH |
| Individual Differences in Two Emotion Regulation Processes: Implications for Affect, Relationships, and Well-Being | Article | 2003 | Journal of Personality and Social Psychology | 7851 | USA | NIH |
| Glide: A New Approach for Rapid, Accurate Docking and Scoring. 1. Method and Assessment of Docking Accuracy | Resource | 2004 | Journal of Medicinal Chemistry | 7775 | USA | NIH |
| Neutrophil Extracellular Traps Kill Bacteria | Article | 2004 | Science | 7735 | Germany | NIH |
| Statistical significance for genomewide studies | Article | 2003 | PNAS | 7684 | USA | None |
| PGC-1α-responsive genes involved in oxidative phosphorylation are coordinately downregulated in human diabetes | Article | 2003 | Nature Genetics | 7474 | USA | HHMI/non-profit/for-profit/Sweden |
| A Modified Poisson Regression Approach to Prospective Studies with Binary Data | Resource | 2004 | American Journal of Epidemiology | 7234 | Canada | Canada |
| Control of regulatory T cell development by the transcription factor Foxp3 | Article | 2003 | Science | 7173 | Japan | Japan |
| Toll-like receptor signalling | Review | 2004 | Nature Reviews Immunology | 7155 | Japan | Japan/non-profit |
| Development of a new Resilience scale: The Connor-Davidson Resilience scale (CD-RISC) | Resource | 2003 | Depression and Anxiety | 7048 | USA | NIH/for-profit |
| MUSCLE: A multiple sequence alignment method with reduced time and space complexity | Resource | 2004 | BMC Bioinformatics | 6960 | USA | Japan |
| The Epidemiology of Major Depressive Disorder: Results from the National Comorbidity Survey Replication (NCS-R) | Article | 2003 | JAMA | 6817 | USA | NIH |
| Inference of population structure using multilocus genotype data: Linked loci and correlated allele frequencies | Article | 2003 | Genetics | 6812 | Germany | NIH |
| A comparison of normalization methods for high density oligonucleotide array data based on variance and bias | Resource | 2003 | Bioinformatics | 6728 | USA | None |
| Influence of life stress on depression: Moderation by a polymorphism in the 5-HTT gene | Article | 2003 | Science | 6536 | UK | NIH |
| Foxp3 programs the development and function of CD4+CD25+ regulatory T cells | Article | 2003 | Nature Immunology | 6509 | USA | NIH/HHMI/non-profit |
| Polyphenols: Food sources and bioavailability | Review | 2004 | American Journal of Clinical Nutrition | 6503 | France | None |
| Overweight, obesity, and mortality from cancer in a prospectively studied cohort of U.S. Adults | Article | 2003 | NEJM | 6472 | USA | None |
| Identification of human brain tumour initiating cells | Article | 2004 | Nature | 6462 | Canada | Canada/non-profit/institution |
| Mild cognitive impairment as a diagnostic entity | Review | 2004 | Journal of Internal Medicine | 6296 | USA | NIH |
| Network biology: Understanding the cell's functional organization | Review | 2004 | Nature Reviews Genetics | 6269 | USA | NIH/NSF/DOE |
| The fourth report on the diagnosis, evaluation, and treatment of high blood pressure in children and adolescents | Consensus | 2004 | Pediatrics | 6202 | USA | NIH |
| Multidimensional Assessment of Emotion Regulation and Dysregulation: Development, Factor Structure, and Initial Validation of the Difficulties in Emotion Regulation Scale | Resource | 2004 | Journal of Psychopathology and Behavioral Assessment | 6099 | USA | Institution |
| MicroRNAs: Small RNAs with a big role in gene regulation | Review | 2004 | Nature Reviews Genetics | 6068 | USA | NIH |
| The mirror-neuron system | Review | 2004 | Annual Review of Neuroscience | 6045 | Italy | None |
| Confidence limits for the indirect effect: Distribution of the product and resampling methods | Resource | 2004 | Multivariate Behavioral Research | 6012 | USA | NIH |
| Normalization of real-time quantitative reverse transcription-PCR data: A model-based variance estimation approach to identify genes suited for | Article | 2004 | Cancer Research | 5955 | Denmark | European |
| normalization, applied to bladder and colon cancer data sets |  |  |  |  |  |  |
| Mass spectrometry-based proteomics | Review | 2003 | Nature | 5925 | USA | NIH/for-profit/Denmark |
| Photodynamic therapy for cancer | Review | 2003 | Nature Reviews<br>Cancer | 5914 | USA | NIH/non-profit |
| Bariatric surgery: A systematic review and meta-analysis | Review | 2004 | JAMA | 5849 | USA | For-profit |
| Targeting HIF-1 for cancer therapy | Review | 2003 | Nature Reviews<br>Cancer | 5789 | USA | None |
| Improved prediction of signal peptides: SignalP 3.0 | Resource | 2004 | Journal of<br>Molecular<br>Biology | 5767 | Denmark | Denmark/for-profit |
| Acute renal failure - definition, outcome measures, animal models, fluid therapy and information technology needs: the Second International Consensus Conference of the Acute Dialysis Quality Initiative (ADQI) Group. | Consensus | 2004 | Critical care<br>(London,<br>England) | 5694 | Australia | None |
| Chronic inflammation in fat plays a crucial role in the development of obesity-related insulin resistance | Article | 2003 | Journal of<br>Clinical<br>Investigation | 5600 | USA | None |
| Markers of inflammation and cardiovascular disease: Application to clinical and public health practice: A statement for healthcare professionals from the centers for disease control and prevention and the American Heart Association | Consensus | 2003 | Circulation | 5586 | USA | CDC/For-profit |
| Bacterial biofilms: From the natural environment to infectious diseases | Review | 2004 | Nature Reviews<br>Microbiology | 5585 | USA | NIH |
| Methodological index for non-randomized studies (Minors): Development and validation of a new instrument | Resource | 2003 | ANZ Journal of<br>Surgery | 5548 | France | None |
| Osteoclast differentiation and activation | Review | 2003 | Nature | 5522 | USA | None |
| DnaSP, DNA polymorphism analyses by the coalescent and other methods | Resource | 2003 | Bioinformatics | 5457 | Spain | Spain |
| ARB: A software environment for sequence data | Resource | 2004 | Nucleic Acids Research | 5455 | Germany | Germany |
| Revised 2003 consensus on diagnostic criteria and long-term health risks related to polycystic ovary syndrome | Consensus | 2004 | Fertility and Sterility | 5445 | Netherland | For-profit/societies |
| Revised 2003 consensus on diagnostic criteria and long-term health risks related to polycystic ovary syndrome | Consensus | 2004 | Human Reproduction | 5398 | Netherlands | For-profit/societies |
| Silver nanoparticles as antimicrobial agent: A case study on E. coli as a model for Gram-negative bacteria | Article | 2004 | Journal of Colloid and Interface Science | 5352 | Croatia | Croatia |
| Functional connectivity in the resting brain: A network analysis of the default mode hypothesis | Article | 2003 | PNAS | 5325 | USA | NIH |
| Epidemiologic classification of human papillomavirus types associated with cervical cancer | Article | 2003 | NEJM | 5297 | Spain | Spain |
| Docetaxel plus prednisone or mitoxantrone plus prednisone for advanced prostate cancer | Article | 2004 | NEJM | 5269 | Canada | For-profit |
| A multigene assay to predict recurrence of tamoxifen-treated, node-negative breast cancer | Article | 2004 | NeEJM | 5252 | USA | NIH/for-profit |
| Preoperative versus postoperative chemoradiotherapy for rectal cancer | Article | 2004 | NEJM | 5249 | Germany | Germany |
| Mindfulness-based interventions in context: Past, present, and future | Review | 2003 | Clinical Psychology: Science and Practice | 5210 | USA | None |
| Implications of recent clinical trials for the National Cholesterol Education Program Adult Treatment Panel III guidelines | Consensus | 2004 | Circulation | 5198 | USA | NIH/societies |
| The international HapMap project | Resource | 2003 | Nature | 5184 | UK | NIH/non-profit/for-profit/international/institutional |
| The chemokine system in diverse forms of macrophage activation and polarization | Review | 2004 | Trends in Immunology | 5175 | Italy | Italy |
| Mechanisms of TGF-β signaling from cell membrane to the nucleus | Review | 2003 | Cell | 5153 | USA | NIH/HHMI |
| Brown Adipose Tissue: Function and Physiological Significance | Review | 2004 | Physiological Reviews | 5152 | Sweden | Sweden |
| The epidemiology of sepsis in the United States from 1979 through 2000 | Article | 2003 | NEJM | 5131 | USA | NIH |
| Alternative activation of macrophages | Review | 2003 | Nature Reviews Immunology | 5121 | UK | UK/non-profit |
| Cardiac-Resynchronization Therapy with or without an Implantable Defibrillator in Advanced Chronic Heart Failure | Article | 2004 | NEJM | 5104 | USA | For-profit |
| Epigenetic regulation of gene expression: How the genome integrates intrinsic and environmental signals | Review | 2003 | Nature Genetics | 5062 | USA | None |
| The gut microbiota as an environmental factor that regulates fat storage | Article | 2004 | PNAS | 5048 | USA | NIH |
| Hazards of heavy metal contamination | Review | 2003 | British Medical Bulletin | 5037 | UK | None |
| 2001 SCCM/ESICM/ACCP/ATS/SIS International Sepsis Definitions Conference | Consensus | 2003 | Critical Care Medicine | 5008 | USA | Societies |
| SIFT: Predicting amino acid changes that affect protein function | Resource | 2003 | Nucleic Acids Research | 4983 | USA | NIH |

**Supplementary Table 2.** 100 cited biomedical papers published in 2013-4

| Title | Type of paper | Year | Source title | Cited | Country | Funding |
| --- | --- | --- | --- | --- | --- | --- |
| Moderated estimation of fold change and dispersion for RNA-seq data with DESeq2 | Resource | 2014 | Genome Biology | 53866 | Germany | NIH/European/institution |
| Trimmomatic: A flexible trimmer for Illumina sequence data | Resource | 2014 | Bioinformatics | 41694 | Germany | Germany |
| MEGA6: Molecular evolutionary genetics analysis version 6.0 | Resource | 2013 | Molecular Biology and Evolution | 36548 | USA | NIH/Japan |
| MAFFT multiple sequence alignment software version 7: Improvements in performance and usability | Resource | 2013 | Molecular Biology and Evolution | 30829 | Japan | Japan |
| STAR: Ultrafast universal RNA-seq aligner | Resource | 2013 | Bioinformatics | 29530 | USA | NIH |
| RAXML version 8: A tool for phylogenetic analysis and post-analysis of large phylogenies | Resource | 2014 | Bioinformatics | 24870 | Germany | Institution |
| The SILVA ribosomal RNA gene database project: Improved data processing and web-based tools | Resource | 2013 | Nucleic Acids Research | 21199 | Germany | Germany |
| FeatureCounts: An efficient general purpose program for assigning sequence reads to genomic features | Resource | 2014 | Bioinformatics | 14943 | USA | Australia |
| Investigation of the freely available easy-to-use software 'EZR' for medical statistics | Resource | 2013 | Bone Marrow Transplantation | 13173 | Japan | None listed |
| UPARSE: Highly accurate OTU sequences from microbial amplicon reads | Resource | 2013 | Nature Methods | 12968 | USA | Non-profit/institution |
| Phyloseq: An R Package for Reproducible Interactive Analysis and Graphics of Microbiome Census Data | Resource | 2013 | PLoS ONE | 12749 | USA | NIH |
| Multiplex genome engineering using CRISPR/Cas systems | Article | 2013 | Science | 12157 | USA | NIH |
| Cancer statistics, 2013 | Reference statistics | 2013 | CA Cancer | 11912 | USA | Societies |
| Cancer statistics, 2014 | Reference statistics | 2014 | CA Cancer | 11514 | USA | Societies |
| Prokka: Rapid prokaryotic genome annotation | Resource | 2014 | Bioinformatics | 11508 | Australia | Institution |
| Integrative analysis of complex cancer genomics and clinical profiles using the cBioPortal | Resource | 2013 | Science Signaling | 11145 | USA | NIH |
| The hallmarks of aging | Review | 2013 | Cell | 10740 | Spain | Multiple international/non-profit/for-profit |
| TopHat2: Accurate alignment of transcriptomes in the presence of insertions, deletions and gene fusions | Resource | 2013 | Genome Biology | 9853 | USA | NIH |
| Global, regional, and national prevalence of overweight and obesity in children and adults during 1980-2013: A systematic analysis for the Global Burden of Disease Study 2013 | Reference statistics | 2014 | Lancet | 9318 | USA | NIH/for-profit/non-profit/international |
| Executive functions | Review | 2013 | Annual Review of Psychology | 8721 | Canada | NIH |
| GSVA: Gene set variation analysis for microarray and RNA-Seq data | Resource | 2013 | BMC Bioinformatics | 8442 | Spain | NIH/Spain |
| Genome engineering using the CRISPR-Cas9 system | Article | 2013 | Nature Protocols | 8184 | USA | NIH/non-profit |
| RNA-guided human genome engineering via Cas9 | Article | 2013 | Science | 7500 | USA | NIH |
| Signatures of mutational processes in human cancer | Article | 2013 | Nature | 7461 | UK | NIH/European |
| Random effects structure for confirmatory hypothesis testing: Keep it maximal | Article | 2013 | Journal of Memory and Language | 7436 | UK | NIH/UK |
| Diet rapidly and reproducibly alters the human gut microbiome | Article | 2014 | Nature | 7344 | USA | NIH |
| NCBI GEO: Archive for functional genomics data sets – Update | Resource | 2013 | Nucleic Acids Research | 7202 | USA | NIH |
| Predictive functional profiling of microbial communities using 16S rRNA marker gene sequences | Article | 2013 | Nature Biotechnology | 7186 | USA | NIH/NSF/ARO/non-profit/Canada |
| The global distribution and burden of dengue | Reference statistics | 2013 | Nature | 7062 | UK | NIH/DHS/non-profit/European |
| Estimating the sample mean and standard deviation from the sample size, median, range and/or interquartile range | Resource | 2014 | BMC Medical Research Methodology | 6971 | Hong Kong | Hong Kong/institution |
| The strengthening the reporting of observational studies in epidemiology (STROBE) statement: Guidelines for reporting observational studies | Consensus | 2014 | International Journal of Surgery | 6842 | Switzerland | European |
| Prevalence of childhood and adult obesity in the United States, 2011-2012 | Reference statistics | 2014 | JAMA | 6701 | USA | CDC |
| Expert consensus document: The international scientific association for probiotics and prebiotics consensus statement on the scope and appropriate use of the term probiotic | Consensus | 2014 | Nature Reviews Gastroenterology and Hepatology | 6695 | USA | Societies |
| 2014 Evidence-based guideline for the management of high blood pressure in adults: Report from the panel members appointed to the Eighth Joint National Committee (JNC 8) | Consensus | 2014 | JAMA | 6601 | USA | NIH |
| Frailty in elderly people | Review | 2013 | Lancet | 6553 | UK | None listed |
| Molecular mechanisms of epithelial-mesenchymal transition | Review | 2014 | Nature Reviews<br>Molecular Cell<br>Biology | 6501 | USA | NIH |
| Natural RNA circles function as efficient microRNA sponges | Article | 2013 | Nature | 6496 | Denmark | Denmark/non-profit |
| Standards for reporting qualitative research: A synthesis of recommendations | Consensus | 2014 | Academic<br>Medicine | 6490 | USA | None listed |
| Using the framework method for the analysis of qualitative data in multi-disciplinary health research | Resource | 2013 | BMC Medical<br>Research<br>Methodology | 6488 | UK | UK |
| Circular RNAs are a large class of animal RNAs with regulatory potency | Article | 2013 | Nature | 6481 | Germany | Germany/institution |
| QUAST: Quality assessment tool for genome assemblies | Resource | 2013 | Bioinformatics | 6456 | Russia | NIH |
| Extracellular vesicles: Exosomes, microvesicles, and friends | Review | 2013 | Journal of Cell<br>Biology | 6455 | France | France/for-profit/institution |
| Better reporting of interventions: Template for intervention description and replication (TIDieR) checklist and guide | Consensus | 2014 | BMJ | 6214 | Australia | UK/Australia/non-profit |
| The International Classification of Headache Disorders, 3rd edition (beta version) | Consensus | 2013 | Cephalalgia | 6190 | USA | NIH/societies/for-profit/institution/Canada |
| Integrative Genomics Viewer (IGV): High-performance genomics data visualization and exploration | Resource | 2013 | Briefings in<br>Bioinformatics | 6170 | USA | NIH |
| Evaluation of general 16S ribosomal RNA gene PCR primers for classical and next-generation sequencing-based diversity studies | Article | 2013 | Nucleic Acids<br>Research | 6068 | Germany | Germany/Austria/European |
| Inferring tumour purity and stromal and immune cell admixture from expression data | Article | 2013 | Nature<br>Communications | 6055 | USA | NIH |
| GROMACS 4.5: A high-throughput and highly parallel open source molecular simulation toolkit | Resource | 2013 | Bioinformatics | 6052 | Sweden | NIH/European/Sweden |
| De novo transcript sequence reconstruction from RNA-seq using the Trinity platform for reference generation and analysis | Resource | 2013 | Nature Protocols | 6018 | USA | NIH/NSF/non-profit/for-profit/international |
| Cancer genome landscapes | Review | 2013 | Science | 6010 | USA | NIH |
| Pilon: An integrated tool for comprehensive microbial variant detection and genome assembly improvement | Resource | 2014 | PLoS ONE | 6001 | USA | NIH |
| Microenvironmental regulation of tumor progression and metastasis | Review | 2013 | Nature Medicine | 5957 | USA | NIH/societies/non-profit/Canada |
| Biological insights from 108 schizophrenia-associated genetic loci | Article | 2014 | Nature | 5906 | UK | NIH/VA/UK/European |
| The cancer genome atlas pan-cancer analysis project | Resource | 2013 | Nature Genetics | 5771 | USA | NIH |
| InterProScan 5: Genome-scale protein function classification | Resource | 2014 | Bioinformatics | 5554 | UK | European |
| Projecting cancer incidence and deaths to 2030: The unexpected burden of thyroid, liver, and pancreas cancers in the united states | Reference statistics | 2014 | Cancer Research | 5482 | USA | Non-profit |
| 3D bioprinting of tissues and organs | Review | 2014 | Nature Biotechnology | 5441 | USA | None listed |
| Content analysis and thematic analysis: Implications for conducting a qualitative descriptive study | Review | 2013 | Nursing and Health Sciences | 5418 | Finland | Institution |
| PD-1 blockade induces responses by inhibiting adaptive immune resistance | Article | 2014 | Nature | 5384 | USA | NIH/HHMI/for-profit/non-profit/societies |
| 2013 ACCF/AHA guideline for the management of heart failure: A report of the American college of cardiology foundation/american heart association task force on practice guidelines | Consensus | 2013 | Journal of the American College of Cardiology | 5380 | USA | Societies |
| Maternal and child undernutrition and overweight in low-income and middle-income countries | Review | 2013 | Lancet | 5374 | USA | Non-profit |
| A 3D map of the human genome at kilobase resolution reveals principles of chromatin looping | Article | 2014 | Cell | 5346 | USA | NIH/NSF/non-profit/institution |
| Angiotensin-neprilysin inhibition versus enalapril in heart failure | Article | 2014 | NEJM | 5325 | USA/UK | For-profit |
| Increased survival in pancreatic cancer with nab-paclitaxel plus gemcitabine | Article | 2013 | NEJM | 5254 | USA | NIH |
| Genome sequence-based species delimitation with confidence intervals and improved distance functions | Article | 2013 | BMC Bioinformatics | 5249 | Germany | Germany/Spain |
| Development of a dual-index sequencing strategy and curation pipeline for analyzing amplicon sequence data on the miseq illumina sequencing platform | Resource | 2013 | Applied and Environmental Microbiology | 5161 | USA | NIH |
| 2012 Revised International Chapel Hill consensus conference nomenclature of vasculitides | Consensus | 2013 | Arthritis and Rheumatism | 5161 | USA | None listed |
| Regulation of ferroptotic cancer cell death by GPX4 | Article | 2014 | Cell | 5127 | USA | NIH |
| Power failure: Why small sample size undermines the reliability of neuroscience | Article | 2013 | Nature Reviews Neuroscience | 5098 | UK | UK |
| Global prevalence of glaucoma and projections of glaucoma burden through 2040: A systematic review and meta-analysis | Reference statistics | 2014 | Ophthalmology | 5070 | Singapore | Singapore |
| Oncology meets immunology: The cancer-immunity cycle | Review | 2013 | Immunity | 5060 | USA | None listed |
| ROS function in redox signaling and oxidative stress | Review | 2014 | Current Biology | 5059 | USA | NIH |
| BEAST 2: A Software Platform for Bayesian Evolutionary Analysis | Resource | 2014 | PLoS Computational Biology | 5032 | New Zealand | NIH |
| Classification of acute pancreatitis - 2012: Revision of the Atlanta classification and definitions by international consensus | Consensus | 2013 | Gut | 4963 | USA | Societies |
| Comprehensive molecular characterization of gastric adenocarcinoma | Article | 2014 | Nature | 4953 | USA | NIH |
| Deciphering key features in protein structures with the new ENDscript server | Resource | 2014 | Nucleic Acids Research | 4933 | France | France |
| The carbohydrate-active enzymes database (CAZy) in 2013 | Resource | 2014 | Nucleic Acids Research | 4892 | France | France |
| The behavior change technique taxonomy (v1) of 93 hierarchically clustered techniques: Building an international consensus for the reporting of behavior change interventions | Consensus | 2013 | Annals of Behavioral Medicine | 4862 | UK | UK |
| Biogenesis, secretion, and intercellular interactions of exosomes and other extracellular vesicles | Review | 2014 | Annual Review of Cell and | 4848 | France | France/non-profit |
|  |  |  | Developmental<br>Biology |  |  |  |
| Pfam: The protein families database | Review | 2014 | Nucleic Acids<br>Research | 4846 | USA | HHMI/European/non-profit |
| Surviving sepsis campaign: International guidelines<br>for management of severe sepsis and septic shock:<br>2012 | Consensus | 2013 | Critical Care<br>Medicine | 4840 | USA | Non-profit |
| Heart Disease and Stroke Statistics - 2014 Update: A<br>report from the American Heart Association | Review | 2014 | Circulation | 4810 | USA | NIH |
| The new frontier of genome engineering with<br>CRISPR-Cas9 | Review | 2014 | Science | 4775 | USA/Germany | NIH/NSF/non-profit |
| Analysis of the structural diversity, substitution<br>patterns, and frequency of nitrogen heterocycles<br>among U.S. FDA approved pharmaceuticals | Article | 2014 | Journal of<br>Medicinal<br>Chemistry | 4603 | USA | NSF |
| Enrichr: Interactive and collaborative HTML5 gene<br>list enrichment analysis tool | Resource | 2013 | BMC<br>Bioinformatics | 4601 | USA | NIH |
| From fastQ data to high-confidence variant calls: The<br>genome analysis toolkit best practices pipeline | Consensus | 2013 | Current<br>Protocols in<br>Bioinformatics | 4600 | USA | NIH |
| Macrophage Activation and Polarization:<br>Nomenclature and Experimental Guidelines | Consensus | 2014 | Immunity | 4574 | USA | NIH/UK/European/non-profit |
| Heart disease and stroke statistics-2013 update: A<br>Report from the American Heart Association | Reference<br>statistics | 2013 | Circulation | 4572 | USA | NIH |
| Diagnosis and classification of diabetes mellitus | Consensus | 2014 | Diabetes Care | 4568 | USA | Societies |
| Toxicity, mechanism and health effects of some heavy<br>metals | Review | 2014 | Interdisciplinary<br>Toxicology | 4563 | India | None listed |
| SPIRIT 2013 statement: Defining standard protocol<br>items for clinical trials | Consensus | 2013 | Annals of<br>Internal<br>Medicine | 4545 | Canada | Canada |
| Global burden of disease attributable to mental and<br>substance use disorders: Findings from the Global<br>Burden of Disease Study 2010 | Reference<br>statistics | 2013 | Lancet | 4534 | Australia | NIH/Australia |
| Ultrasensitive fluorescent proteins for imaging<br>neuronal activity | Article | 2013 | Nature | 4512 | USA | NIH/Switzerland/European |
| A general framework for estimating the relative pathogenicity of human genetic variants | Article | 2014 | Nature Genetics | 4510 | USA | NIH |
| Regulation of microRNA biogenesis | Review | 2014 | Nature Reviews<br>Molecular Cell<br>Biology | 4458 | Korea | Korea |
| Ultrasensitive photodetectors based on monolayer MoS 2 | Article | 2013 | Nature<br>Nanotechnology | 4446 | Switzerland | Switzerland/European |
| Chimeric antigen receptor T cells for sustained remissions in leukemia | Article | 2014 | NEJM | 4414 | USA | NIH/for-profit/non-profit/institution |
| Development and applications of CRISPR-Cas9 for genome engineering | Review | 2014 | Cell | 4403 | USA | NIH/NSF/non-profit |
| Cancer incidence and mortality patterns in Europe: Estimates for 40 countries in 2012 | Reference statistics | 2013 | European Journal<br>of Cancer | 4381 | France | European/IARC |
| Lipid peroxidation: Production, metabolism, and signaling mechanisms of malondialdehyde and 4-hydroxy-2-nonenal | Review | 2014 | Oxidative<br>Medicine and<br>Cellular<br>Longevity | 4376 | Spain | Spain |

**Supplementary Table 3.**
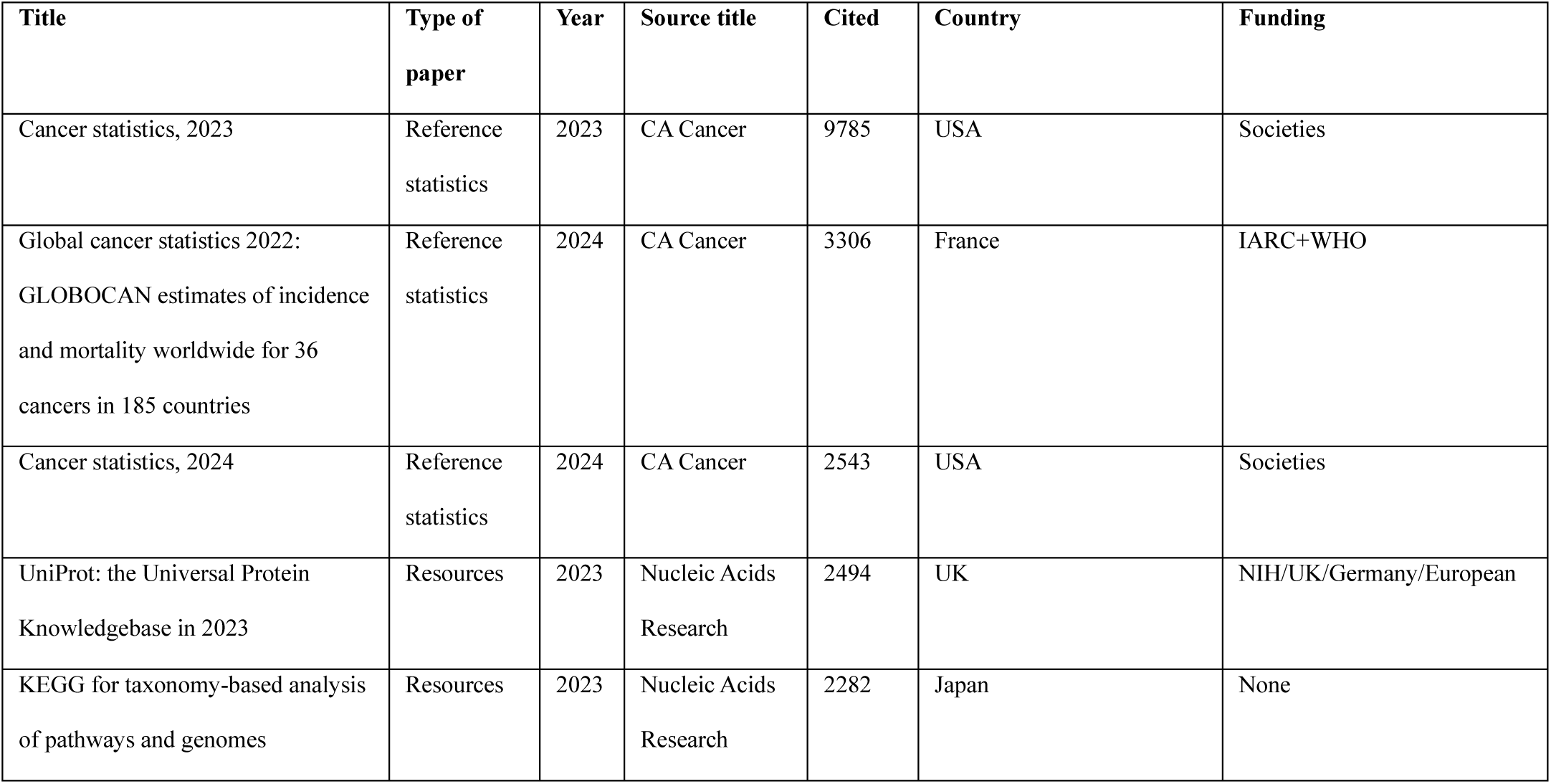
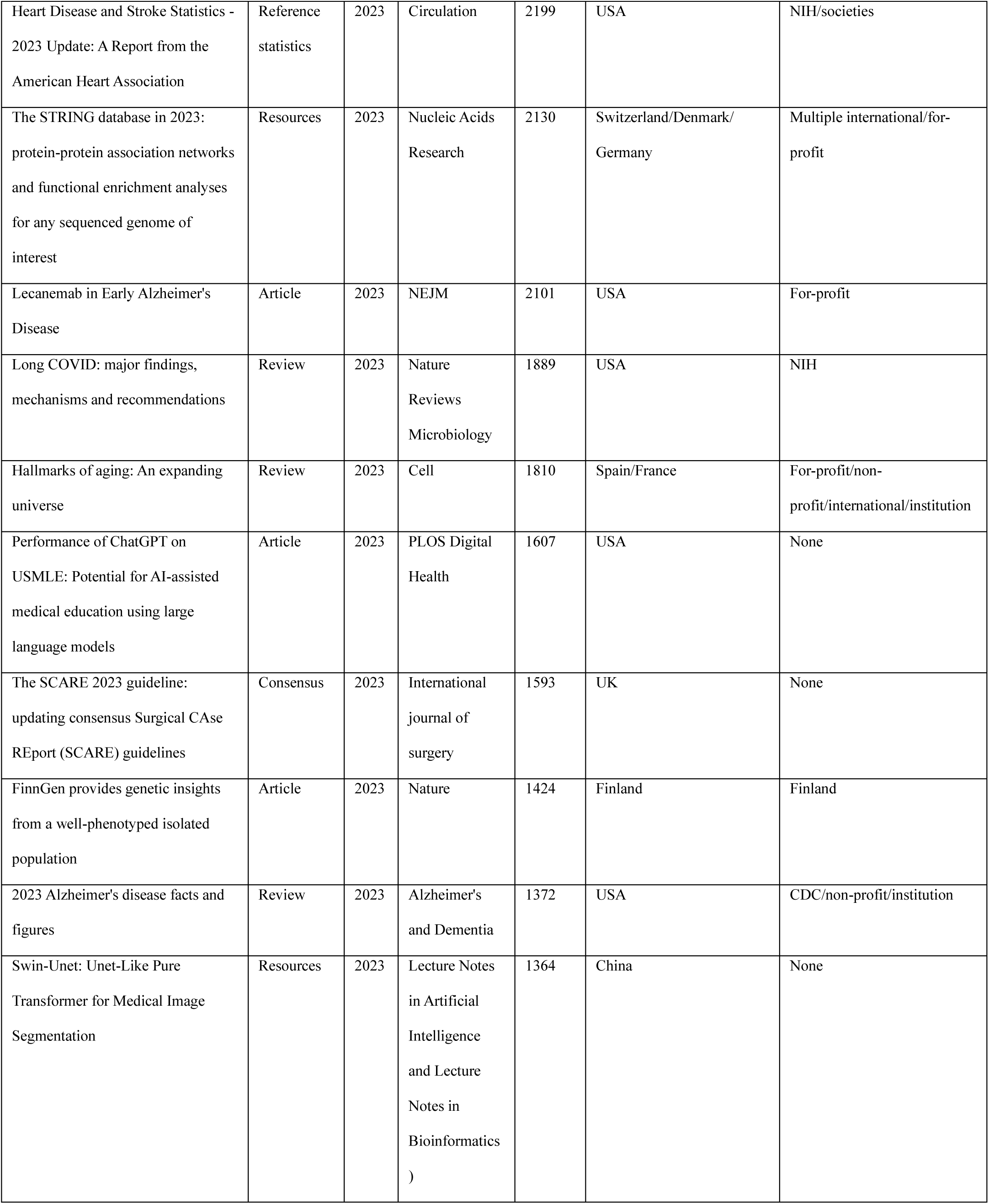
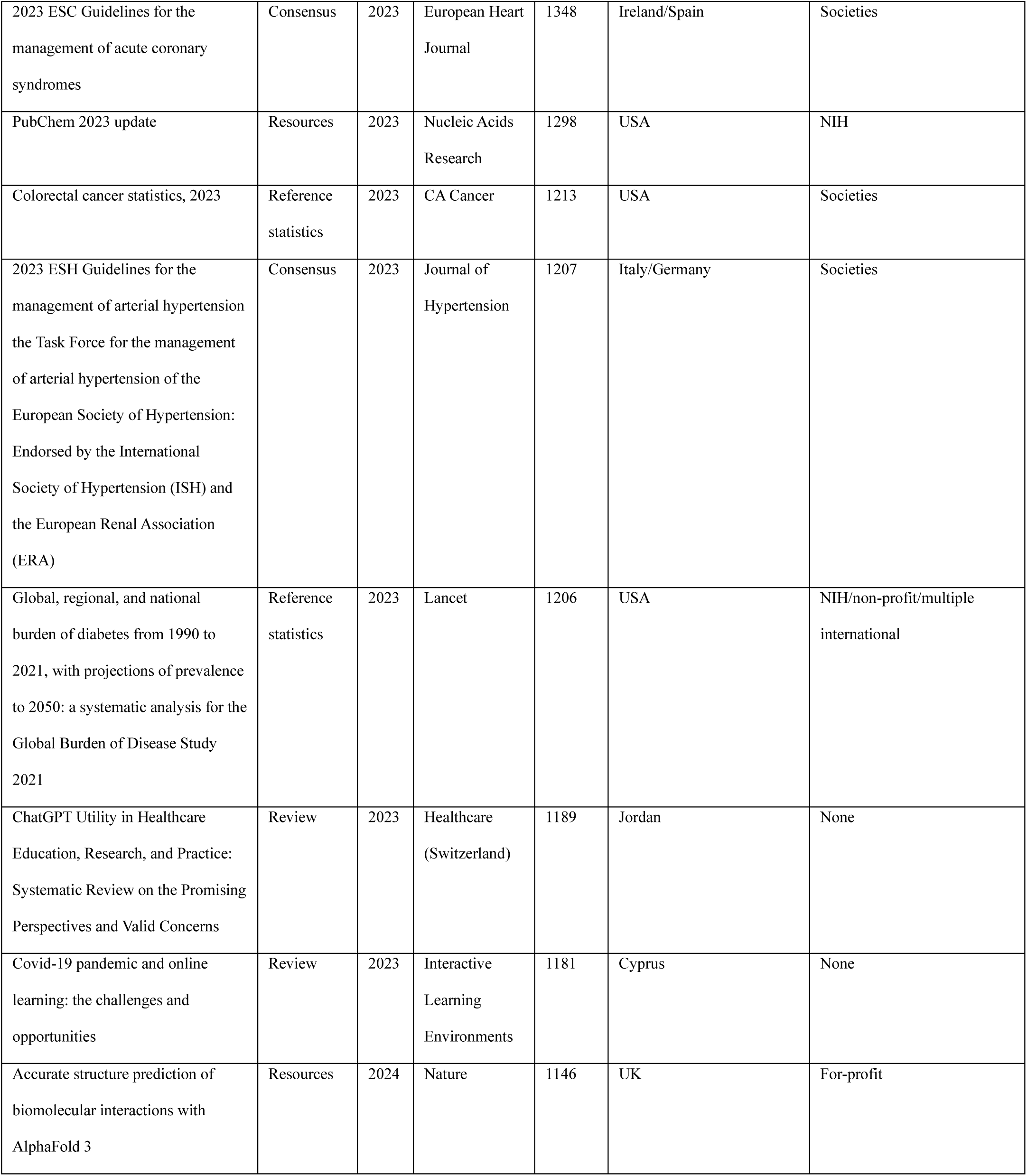
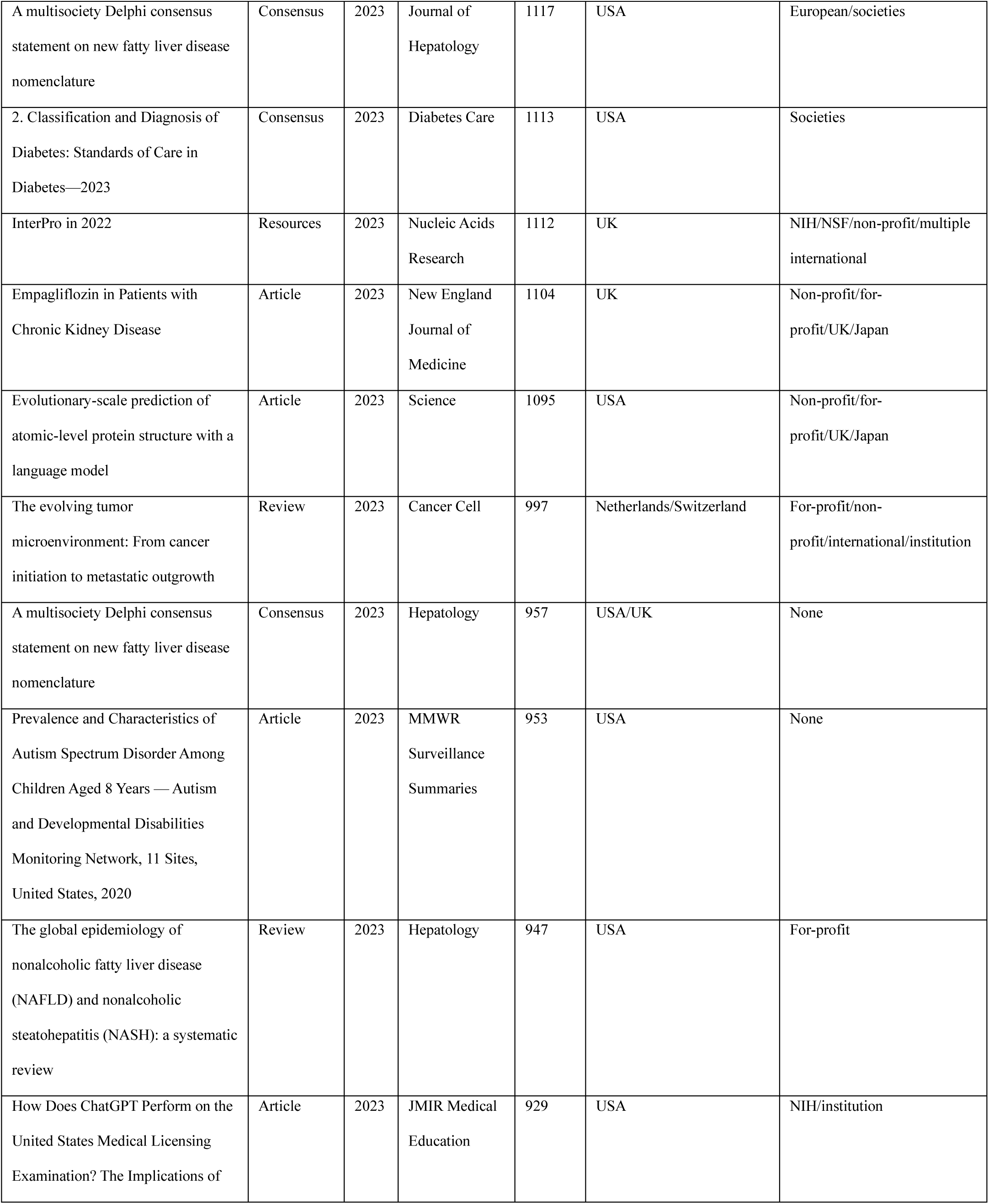
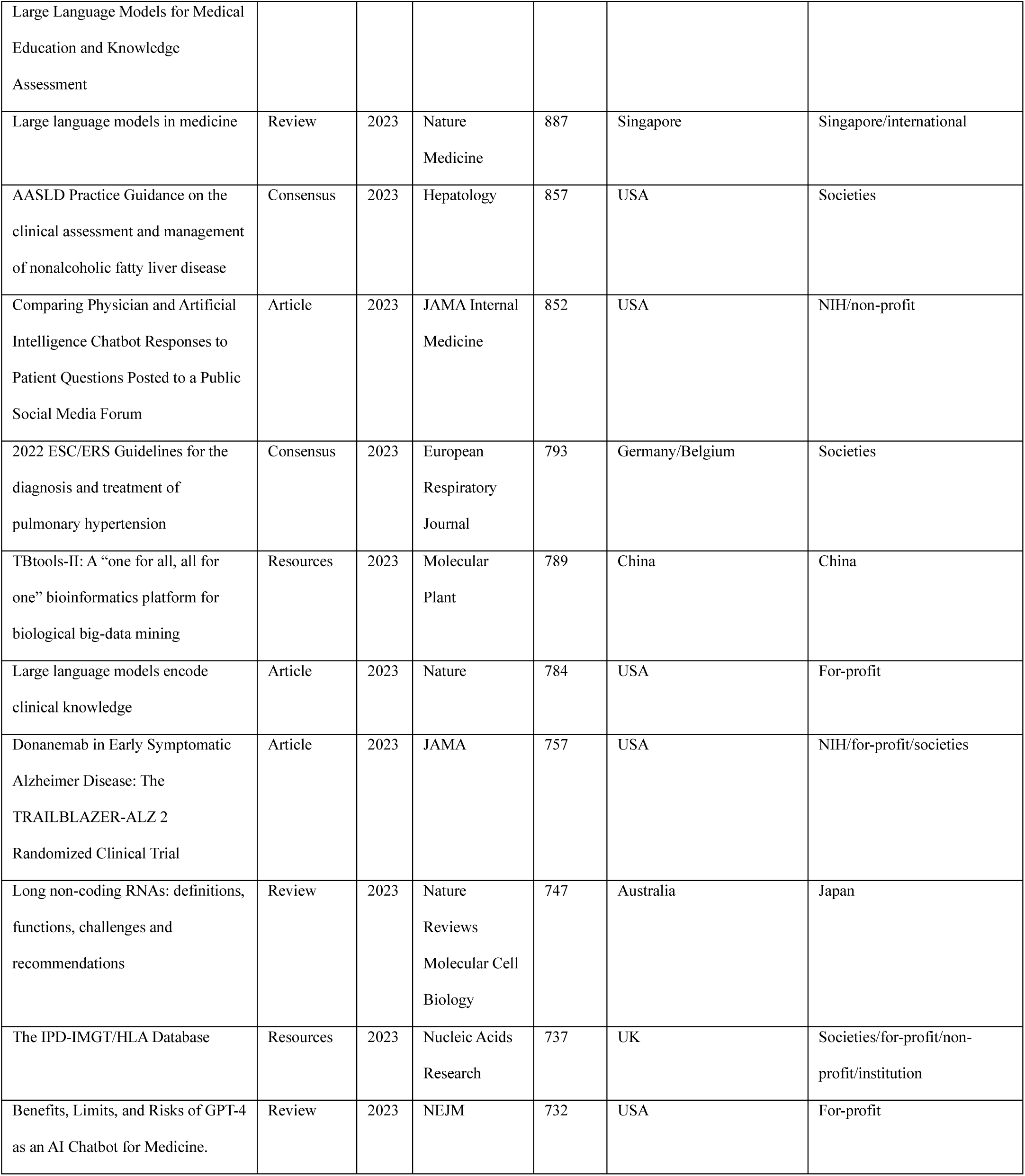
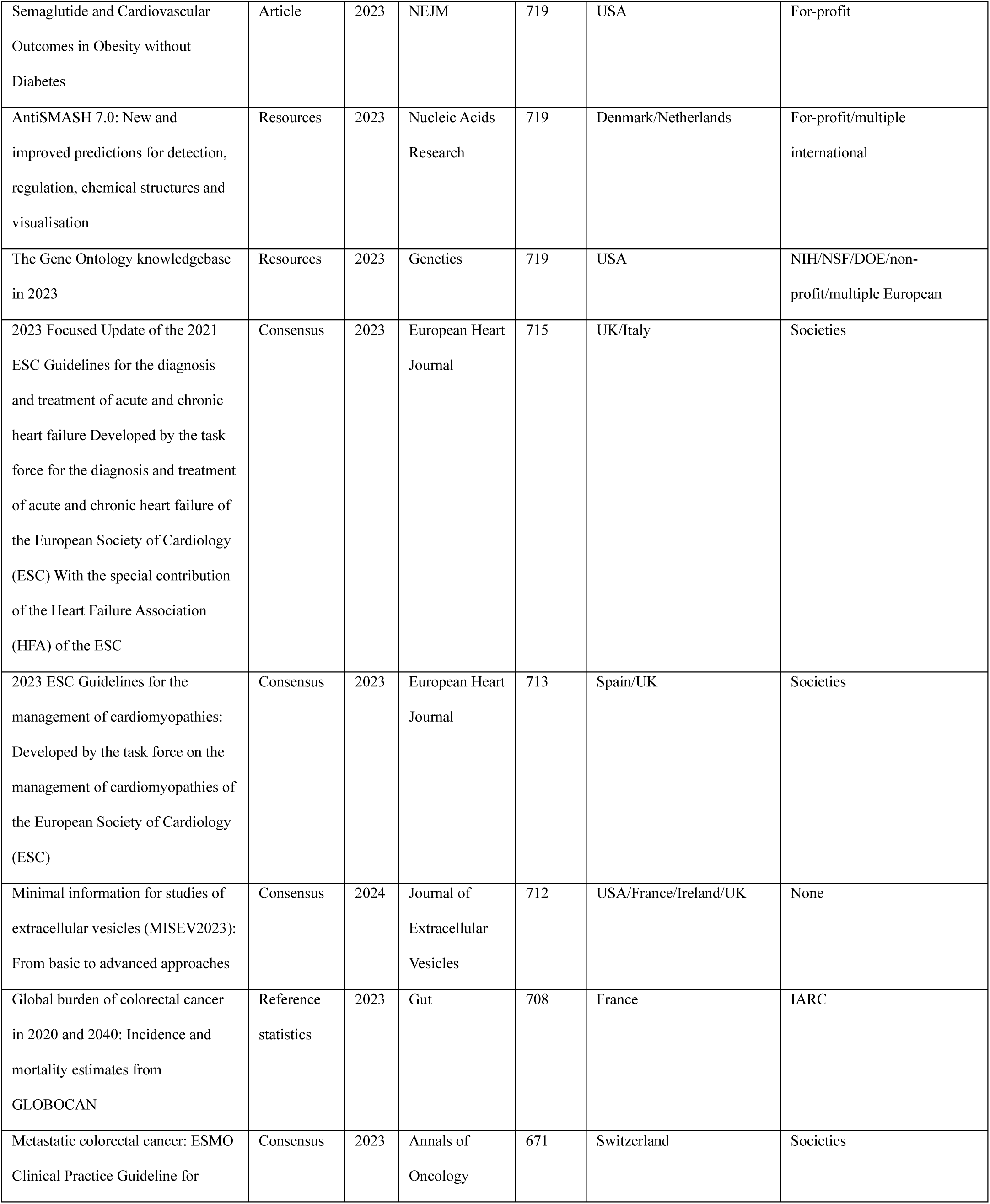
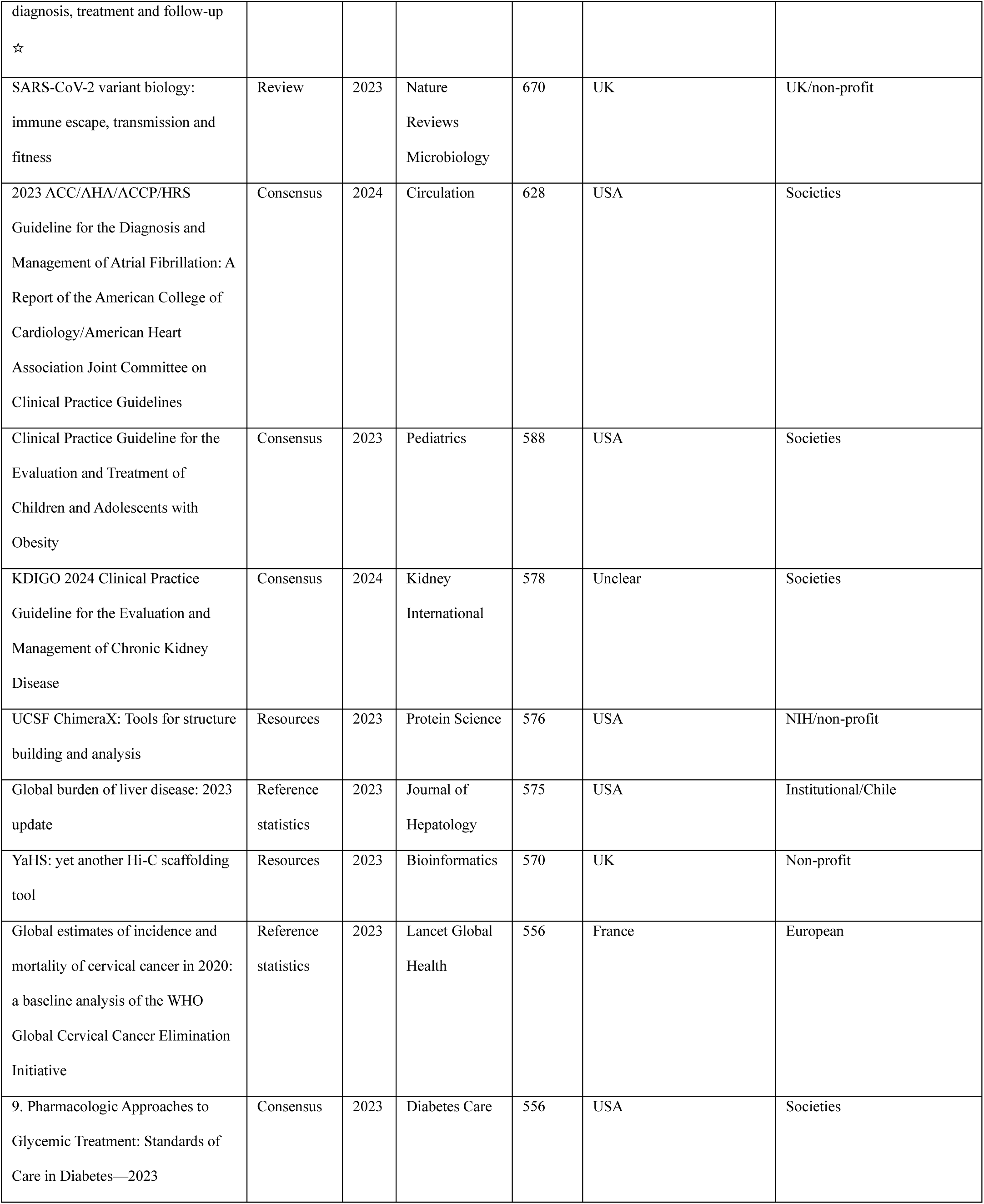
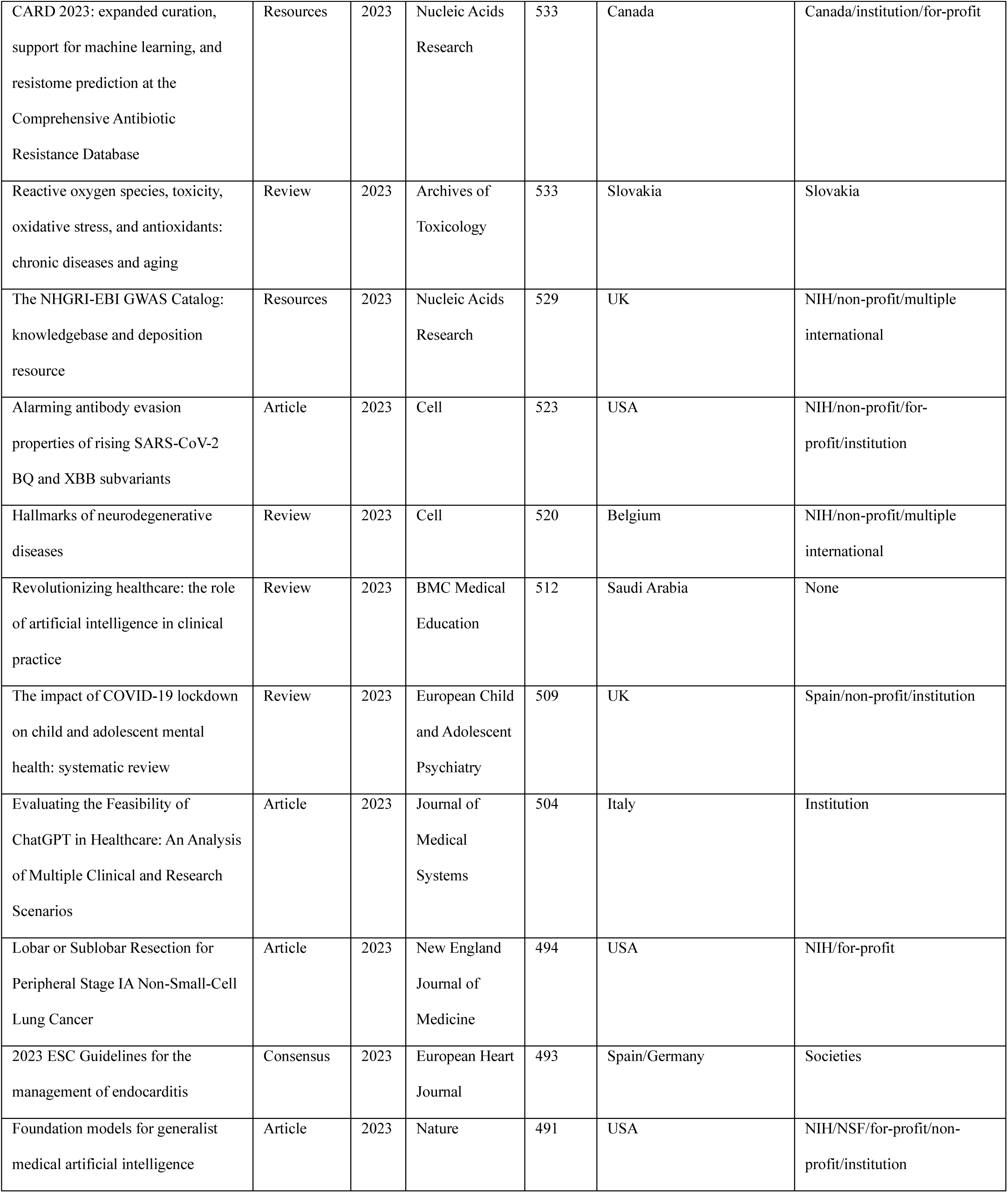
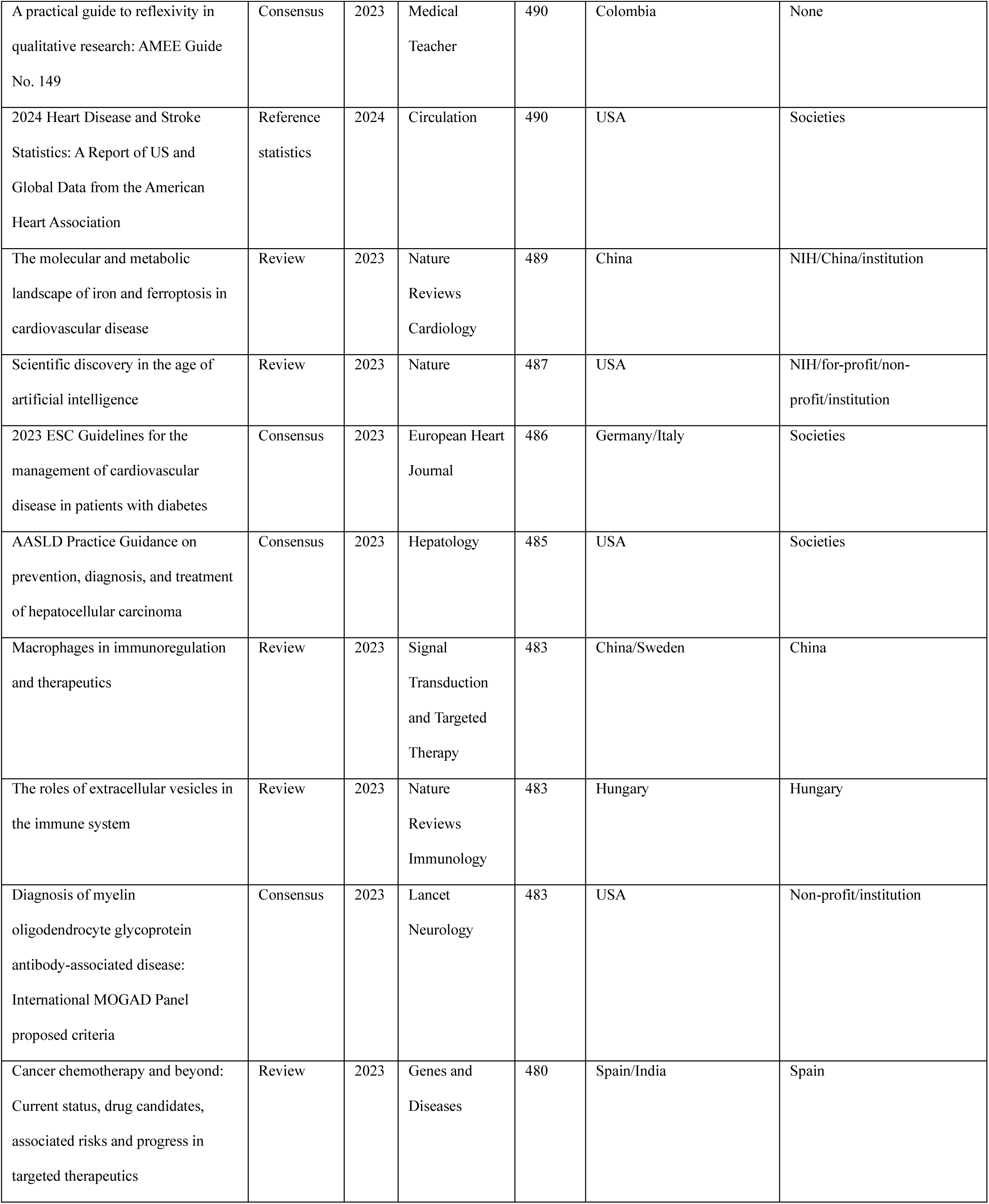
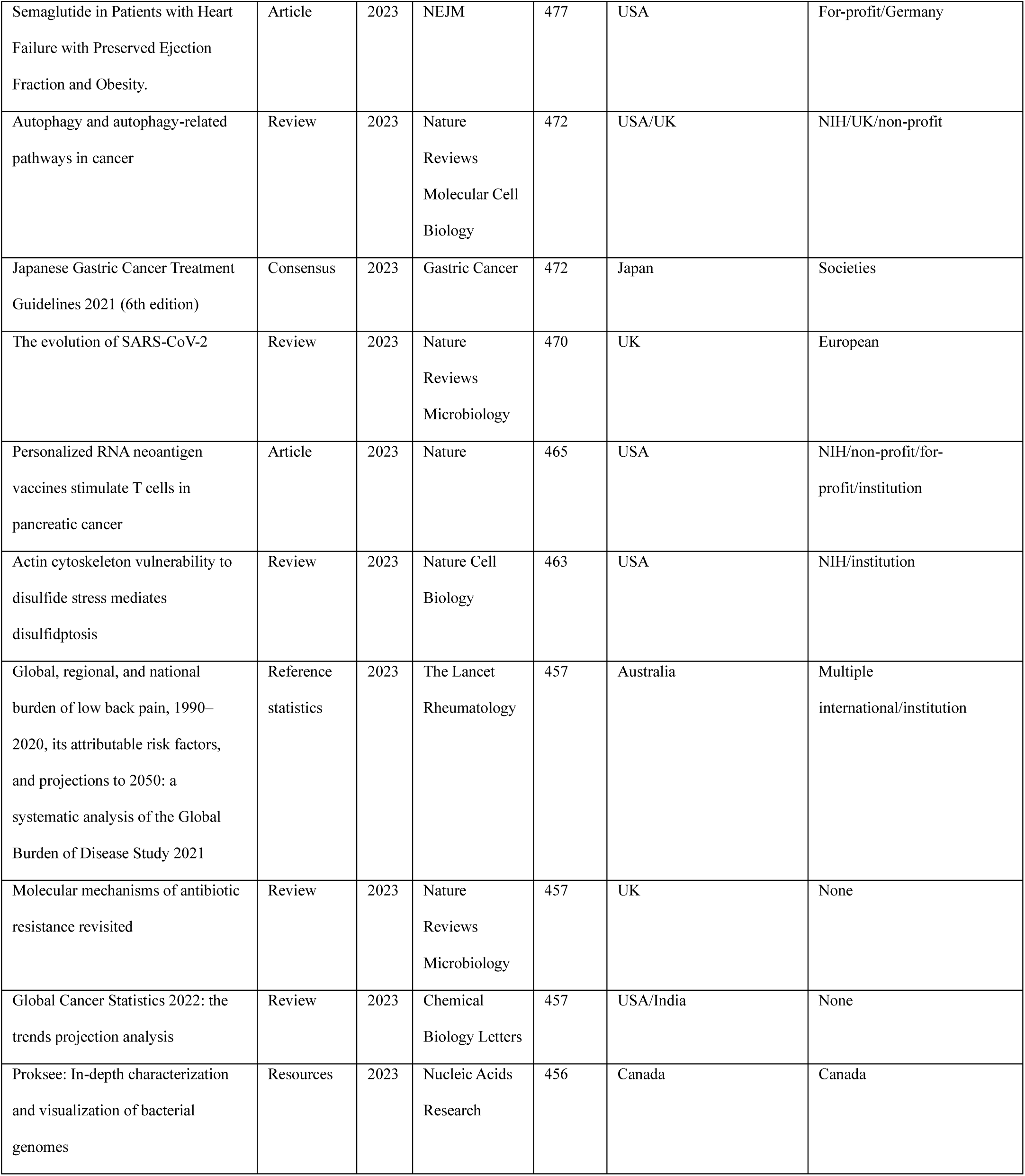
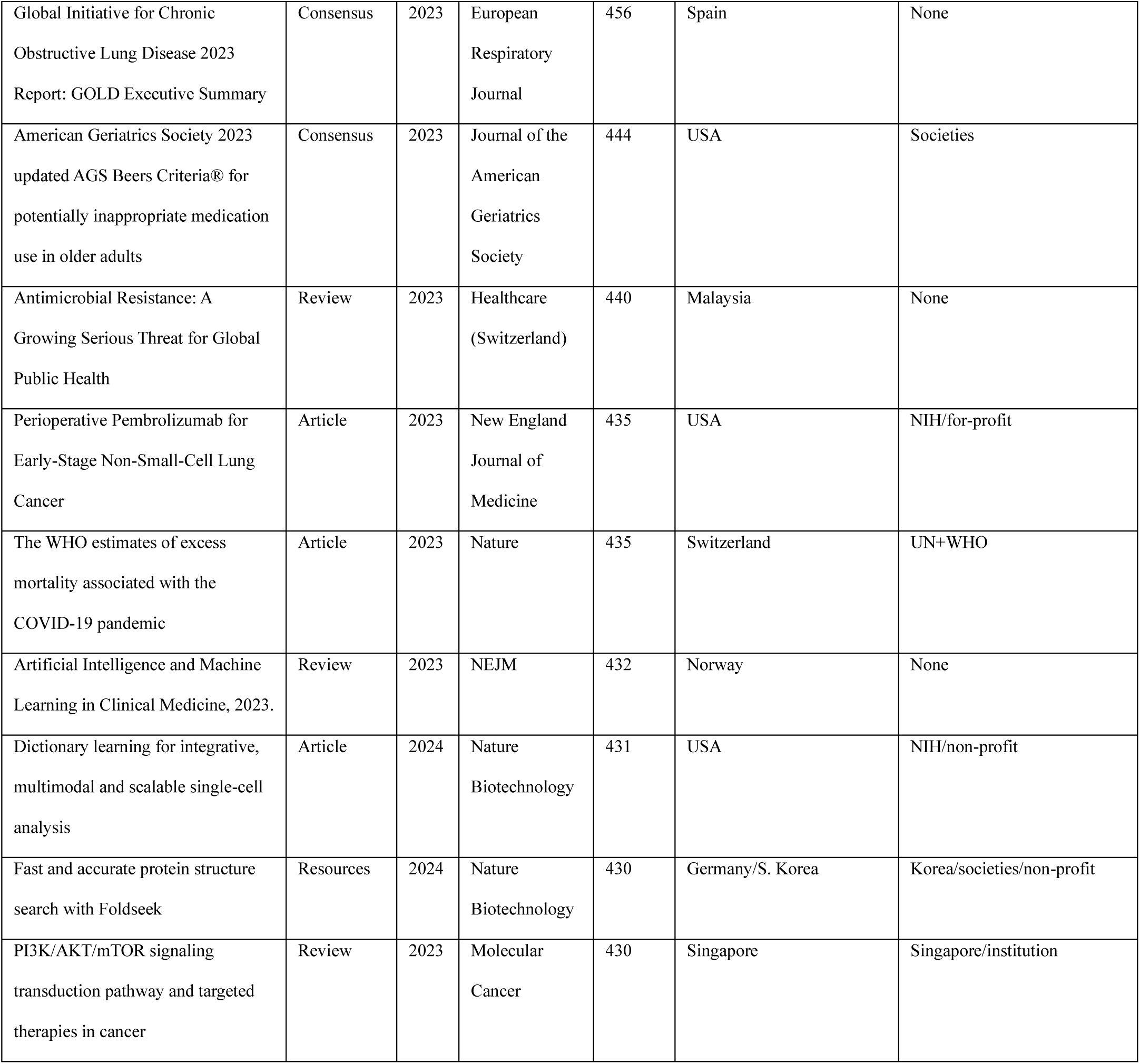
100 top cited biomedical papers published in 2023-4

